# Evolution of a globally unique SARS-CoV-2 Spike E484T monoclonal antibody escape mutation in a persistently infected, immunocompromised individual

**DOI:** 10.1101/2022.04.11.22272784

**Authors:** Peter J. Halfmann, Nicholas R. Minor, Luis A. Haddock, Robert Maddox, Gage K. Moreno, Katarina M. Braun, David A. Baker, Kasen K. Riemersa, Ankur Prasad, Kirsten J. Alman, Matthew C. Lambert, Kelsey Florek, Allen Bateman, Ryan Westergaard, Nasia Safdar, Dave R. Andes, Yoshihiro Kawaoka, Madiha Fida, Joseph D. Yao, Thomas C. Friedrich, David H. O’Connor

## Abstract

Prolonged infections in immunocompromised individuals may be a source for novel SARS-CoV-2 variants, particularly when both the immune system and antiviral therapy fail to clear the infection, thereby promoting adaptation. Here we describe an approximately 16-month case of SARS-CoV-2 infection in an immunocompromised individual. Following monotherapy with the monoclonal antibody Bamlanivimab, the individual’s virus was resistant to this antibody via a globally unique Spike amino acid variant (E484T) that evolved from E484A earlier in infection. With the emergence and spread of the Omicron Variant of Concern, which also contains Spike E484A, E484T may arise again as an antibody-resistant derivative of E484A.

## Introduction

The host immune response typically clears SARS-CoV-2 infections by 15 to 25 days ^1–4^, and this short duration of active virus replication limits within-host evolution. In contrast, infections in immunocompromised individuals can be prolonged, with the most prolonged previously documented infection lasting 333 days ^5^. Prolonged infections may facilitate SARS-CoV-2 evolution by providing time for within-host selection to act, potentially driving the emergence of immunologic escape variants (reviewed in ^6^).

Spike residue E484 lies in the receptor-binding domain (RBD), which is the target for approximately 90% of neutralizing antibodies ^7^. Class 2 Spike neutralizing antibodies, as defined in Barnes et al. 2020, are particularly sensitive to Spike residue 484 substitutions ^8^. Class 2 neutralizing antibodies form a hydrogen bond network with Spike residue 484, which is highly conserved across class 2 antibodies. A recent study exposed a VSV-eGFP-SARS-CoV-2-S chimeric virus to 19 neutralizing monoclonal antibodies (mAbs) and found that substitutions appeared at E484 more frequently than at any other residue. E484A, E484D, E484G, and E484K all reduce the neutralization activity of antibodies in convalescent plasma ^9^. Spike E484K is the best studied of these variants; the substitution was found to reduce susceptibility to the monoclonal antibody Bamlanivimab by more than 2,360-fold ^10^. E484K is found in the Beta (B.1.351) and Gamma (P.1) Variants of Concern as well as the B.1.617.1 and P.2 lineages, all of which were commonly circulating in the spring of 2020. This prompted the US Food and Drug Administration to revoke Emergency Use Authorization for Bamlanivimab on April 16, 202110.

A second variant at this residue, Spike E484A, is found in the highly transmissible Omicron (B.1.1.529) Variant of Concern. Prolonged infection in an immunocompromised individual has been hypothesized as the source of Omicron ^11^. Providing further support for this hypothesis, E484A variants have also arisen in immunocompromised individuals with prolonged infection ^11,12^. SARS-CoV-2 culture in the presence of monoclonal antibodies can also select E484A *in vitro* ^13^. E484A was transiently detected in one patient receiving Bamlanivimab monotherapy, but the impact of this mutation on Bamlanivimab recognition was not directly tested ^14^.

Here, we report an ongoing, 16-month-long SARS-CoV-2 infection in an immunocompromised patient. Following unsuccessful Bamlanivimab monotherapy, a number of substitutions were detected in the consensus sequence of the infecting virus, including one encoding Spike E484T, a substitution that has not been reported previously in public SARS-CoV-2 sequence databases. Virus isolated from this patient encoding Spike E484T was highly resistant to Bamlanivimab, but remained susceptible to three other commercial monoclonal antibodies. These findings highlight the necessity of frequent testing, viral sequencing, and proactive treatment for prolonged SARS-CoV-2 infections occurring in immuno-compromised individuals.

## Results

### Case description

The patient described in this case is in their 50s and is immunocompromised. In Fall 2020, the patient reported sinus congestion and pain breathing to their primary care provider, and a nasopharyngeal swab specimen first tested PCR-positive for SARS-CoV-2. The patient’s symptoms worsened into fever, shortness of breath, and body fatigue by ∼two months after their initial diagnosis. The patient was admitted to a hospital emergency department, where physicians found evidence for organizing pneumonia and alveolar damage, and no evidence of bacterial or fungal infection. Despite hypoxia and recurrent fevers, the patient’s blood oxygen saturation remained in the healthy range above 90 mmHg, and they were sent home. There, the patient started a course of dexamethasone. Their fevers improved, but their hypoxia worsened, requiring supplemental oxygen via nasal cannula during sleep and activity. The patient’s blood oxygen saturation continued to fall into the 70s, requiring placement on a bilevel positive airway pressure (BiPAP) ventilator to bring oxygenation back into the 90s.

About four months post-diagnosis, the patient was admitted to a hospital Intensive Care Unit (ICU) and immediately intubated. The patient was extubated less than a week later, though they were unable to wean off supplemental oxygen via nasal cannula. A nasopharyngeal swab specimen from the patient once again tested PCR-positive for SARS-CoV-2, which prompted the ICU physicians to administer two courses of inpatient remdesivir, two units of convalescent plasma, and an aggressive steroid regimen. The physicians also requested compassionate use of monoclonal antibodies from Eli Lilly and Regeneron, though at the time, both companies denied their requests. Yet another nasopharyngeal swab PCR test came back positive for SARS-CoV-2. The patient received intravenous immunoglobulin, and less than a week later the patient was discharged on a steroid regimen and supplemental oxygen via nasal cannula.

Around six months post-diagnosis, the physicians submitted another request for compassionate use of a monoclonal antibody therapy to Eli Lilly. This time, the company approved their request, and the patient received 700mg of intravenous Bamlanivimab. More than a month later, the patient reported worsening shortness of breath and low-grade fever, and chest CT scans again showed organizing pneumonia. Additional nasopharyngeal swabs came back positive, though without any major changes in symptoms and pulmonary function. Eventually, at almost 13 months post-diagnosis, the patient’s cough and difficulty breathing worsened again, at which point they sought a second opinion from another facility. After another PCR-positive nasopharyngeal swab, the physicians administered three infusions of high-titer convalescent plasma as well as five days of IV remdesivir. Bacteria and fungal cultures remained negative, as was a bronchoalveolar lavage sample for SARS-CoV-2. Still, the patient’s nasopharyngeal swabs remained PCR-positive for SARS-CoV-2 infection.

At about 14 months post-diagnosis, the patient was started on three days of outpatient VaxPlasma, a high-titer donor blood plasma from vaccinated individuals. The infusion boosted the patient’s anti-spike protein antibody titers by more than twofold, but shortly thereafter, nasopharyngeal swabs from the patient remained PCR-positive. Low blood oxygen, requiring two to three liters per minute of supplemental oxygen, and labored breathing at rest persisted during that period. At 15 months post-diagnosis, a transbronchial upper left lung lobe biopsy tested PCR-positive for SARS-CoV-2 RNA. At 16 months post-diagnosis, the patient’s nasopharyngeal swab again tested PCR-positive, and subsequently received another five-day course of IV remdesivir and another three infusions of VaxPlasma. Nonetheless, another nasopharyngeal swab tested positive, though with the highest Ct (indicating the lowest SARS-CoV-2 RNA copy number) of their infection (Supplemental Table 1).

At the time of this writing, it remains unclear whether the patient is experiencing symptoms from a persistent, actively replicating viral infection, or is suffering from post-COVID lung damage with persistent viral excretion in the respiratory tract.

Note that the above case history has been abbreviated, with certain delails withheld at the request of medRxiv. The full case description and timeline is available upon reasonable request to the corresponding author.

### Prolonged detection of SARS-CoV-2 by RT-qPCR and virus culture

Ct values were below 27 in the majority of nasopharyngeal swab PCR tests, indicating active viral replication and high viral loads (Figure 1A). At nearly ten months post-diagnosis, infectious viruses from a nasopharyngeal swab were successfully cultured *in vitro*. Additionally, viral isolates from the immunocompromised individual were sequenced at twelve timepoints, beginning at ∼4 months post-diagnosis. We found that consensus sequences from each of these timepoints are more closely related to one another than any other sequences in public viral sequence repositories and are all Pango lineage B.1.2, consistent with a prolonged infection rather than reinfection (Figure 2B). To our knowledge, this presumptive 16-month ongoing infection is the most prolonged SARS-CoV-2 infection reported to date, and it highlights the need for an arsenal of medical interventions in persistently infected immuno-compromised individuals. Additionally, the virus described in this case accumulated enough mutations within its host to roughly match the global SARS-CoV-2 mutation rate for the whole of the pandemic (see Supplemental Figure 2).

**Figure 01.**
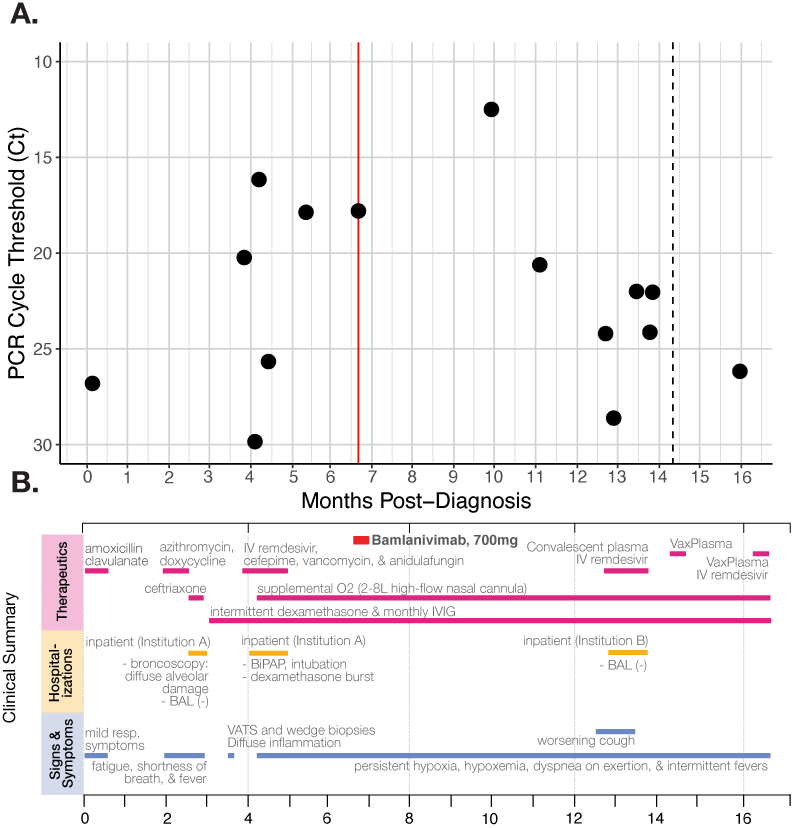
A) RT-qPCR cycle threshold values for patient nasopharyngeal swab specimens collected over almost 16 months of follow-up. The red line marks when the patient received the Bamlanivimab intravenous monoclonal antibody treatment (700mg). The dotted line in month 14 indicates a positive PCR test but no available Ct value. Note that Ct values are inverted to correspond with the inverse relationship between PCR Ct threshold value and and nasal RNA copy number. **B) Timeline of clinically relevant information** for the patient’s chronic infection, including symptoms, therapeutics, and hospitalizations.

**Figure 02.**
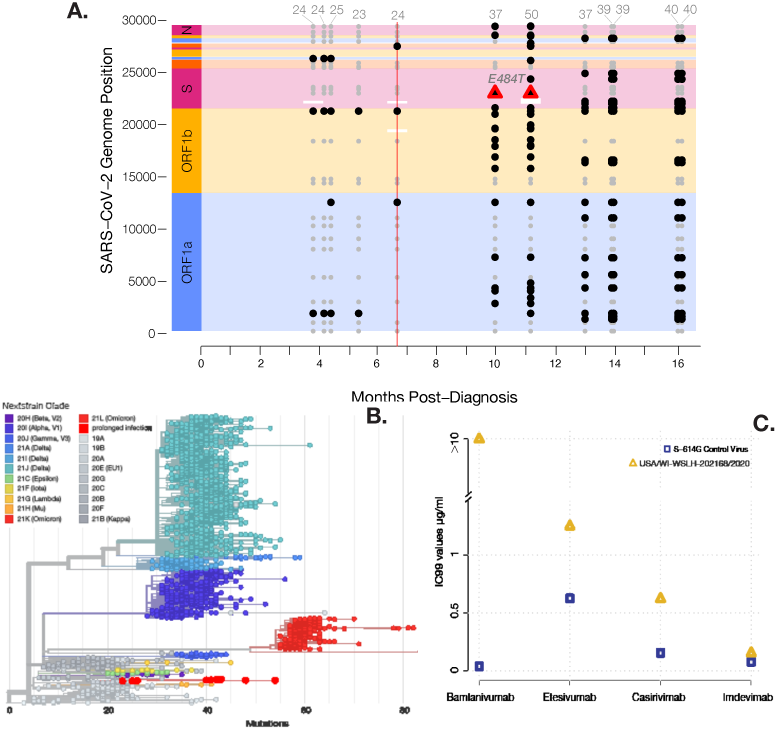
A) Consensus-level mutations (present in >50% of reads) across the SARS-CoV-2 genome throughout infection of an immunocompromised individual. Gray points represent consensus-level mutations that persisted throughout all sampled timepoints and may have been inherited from the infecting lineage. Black points represent consensus-level mutations that emerged or disappeared during the chronic infection. The vertical red line represents Bamlanivimab intravenous mAb treatment (700mg). Amplicon dropouts are depicted as white horizontal bars. Consensus-level E484T mutations in Spike are shown as red triangles. Among the mutations depicted here, three are noncoding, 18 are synonymous, and 44 are nonsynonymous. **B) Globally subsampled SARS-CoV-2 phylogeny with chronically infected patient’s twelve virus sequences**. Patient sequences are indicated with larger red dots toward the bottom of the tree. The patient’s virus descends from an ancestor in the lineage B.1.2. The phylogeny was constructed using UCSC UShER (https://genome.ucsc.edu/cgi-bin/hgPhyloPlace). **C) The concentration of four commercially available antibody treatments required to neutralize 99% of the chronically infected patient’s SARS-CoV-2**. While Etesivumab (Eli Lilly), Casirivimab (Regeneron), and Imdevimab (Regeneron) were able to neutralize virus near or below 1 µg/ml, Bamlanivimab was unable to neutralize the virus at concentrations as high as 10 µg/ml, indicating antibody escape after the previous Bamlanivimab treatment.

A single, prolonged infection is the most parsimonious explanation for these findings. However, a household member tested positive for SARS-CoV-2 about three months post-diagnosis, and it is theoretically possible that the immunocompromised individual cleared their infection between the previously confirmed PCR test on day 0 and was reinfected by their household contact. Unfortunately, no samples from this household member are available to investigate this possibility.

### Emergence of the globally unique Spike E484T and other mutations in response to Bamlanivimab

At the earliest timepoint for which sequencing data is available, taken about four months into the infection, we detected 24 mutations that separated the patient’s virus from Wuhan-1. This number gradually increased to 40 mutations by our final sequencing timepoint. Early mutations included Envelope T30I, one of the most frequently arising mutations immunocompromised individuals ^15^, though this mutation was lost by approximately five months post-diagnosis. Also in our earliest sequencing time-point, we detected a point mutation in Spike codon 484 (GAA>G**C**A), which changed the amino acid glutamic acid (E) to alanine (A). This Spike E484A mutation persisted in the virus consensus sequence for five further timepoints, at least to eight months post-diagnosis. At that point, the patient received the Bamlanivimab monoclonal antibody therapy, and their virus was subsequently not sequenced for more than three months; there was no additional testing or sequencing during this interval. At the next point at which their virus was sequenced, we observed a cumulative total of 37 consensus mutations from the Wuhan-1 sequence, eleven of which had risen together from sub-consensus variants detected at previous timepoints. One of the eleven that arose together was an additional, novel point mutation in the Spike codon 484 (G**C**A>**AC**A), which changed the amino acid residue to threonine (T). As of February 2022, Spike E484T has not been documented in any other SARS-CoV-2 samples. The substitution remained in our next sequencing timepoint at 11 months post-diagnosis. However, by the following timepoint approximately two months later, the nucleotide mutation that caused E484T had returned to a frequency far below consensus, as did ten mutations that rose with it. The virus consensus sequence at ∼13 months post-diagnosis, Spike codon 484 had reverted to E484A (Supplemental Figure 1).

We next examined whether the individual’s virus harboring E484T escaped monoclonal antibody recognition. We cultured the virus from about ten months post-diagnosis, when we detected twenty-five new consensus mutations including Spike E484T (Figure 2A), and performed a neutralization assay with four commercially available antibodies (Figure 2C). Eli Lilly’s Etesivumab, as well as Regeneron’s Casirivimab and Imdevimab, neutralized the virus at relatively similar concentrations. 99% of cell death was prevented with 1.25 µg/mL of Etesivumab, 0.62 µg/mL of Casirivimab, and 0.16 µg/mL of Imdevimab. However, Eli Lilly’s Bamlanivimab could not neutralize the virus at concentrations as high as 10 µg/mL, indicating that Spike E484T is a Bamlanivimab escape variant.

## Discussion

The degree of genetic evolution we document in this patient’s virus highlights the potential role of immunocompromised individuals in generating and propagating novel variants. We also describe the first case of a globally unique Spike E484T variant, which arose after treatment with the Bamlanivimab monotherapy. While E484T appears to be novel for SARS-CoV-2, the bat betacoronavirus RaTG13, one of the most similar coronaviruses to SARS-CoV-2 in terms of nucleotide identity, also bears a threonine at Spike codon 484. This suggests that E484T is a variant that can be tolerated in betacoronaviruses with similar Spike structure to SARS-CoV-2, though the RaTG13 Spike has noteworthy differences, including lower stability in its pre-furin-cleaved form ^16^. Additionally, *in silico* E484T mutants of the SARS-CoV-2 Spike protein showed increased transduction of mouse ACE2 receptors, the key for viral entry into the cell, by more than 16-fold ^17^.

In this case, it is unfortunate–though certainly not unprecedented–that residual specimens from this patient’s earliest SARS-CoV-2 tests are not available for retrospective analyses. Given the volume of specimen required for RT-qPCR, most providers discard residual nasopharyngeal swab specimens after testing. However, this case highlights the potential value of archiving test-positive specimens from immunocompromised individuals. Without sequence data from earlier in the infection, it is impossible to determine conclusively whether the E484A variant arose *de novo* in this individual, or if they were infected with a lineage harboring E484A. However, similar studies of prolonged SARS-CoV-2 infections in immunocompromised individuals ^12,18–20^, the lack of Spike E484A in the ancestral B.1.2 lineage, and the lack of other E484A sequences in Wisconsin during late 2020 indicate that this variant likely arose *de novo* in the months following initial infection. A national or global registry to collect samples from individuals with prolonged infection and to study these individuals in greater numbers than can be accrued at any one clinical site, akin to the International HIV Controllers Study, may be warranted.

Although data are limited in the county where this individual lives, surveillance sequencing suggests that there was no onward spread of the individual’s virus, notwithstanding the household case of unknown provenance. This lack of spread is particularly consequential for the Bamlanivimab-evading virus, which bore the novel Spike E484T along with a cumulative 36 other mutations. That E484T is unique to this individual raises the possibility that E484T, and the other mutations that emerged and disappeared along with it (Supplemental Figure 1), has an associated fitness cost.

Another, non-mutually exclusive explanation for the lack of onward spread is that the patient’s collaboration with state and local public health officials has been successful at managing their infection, especially after Spike E484T was detected, and indeed it is to their credit that this case report involves only one individual. That individual has remained isolated at home for the majority of their 16-month, ongoing infection, and has cooperated with ongoing monitoring efforts by state public health officials. While this is only one case, it foreshadows the difficult ethical questions that come up in any case of prolonged infection in an immunocompromised individual, of which there are millions in the United States ^21^. If these individuals are at a higher risk of prolonged SARS-CoV-2 infections, and these infections pose outsized risks for the emergence of immune escape variants, what responsibility do communities, public health agencies, and policy-makers have to coordinate with these individuals and prevent onward spread? If immunodeficiency is a risk factor for prolonged SARS-CoV-2 shedding and the emergence of worrisome variants, does this create a risk of stigmatizing immunosuppression?

What support is available to these individuals in the event that they cannot maintain employment or afford healthcare? How do therapeutics intended to manage symptoms—or the lack thereof—influence the virus’s evolution? These questions echo previous controversies, such as the isolation of individuals with highly drug-resistant tuberculosis ^22^, but may be amplified in the United States by the high incidence of SARS-CoV-2 and the large number of immunosuppressed/immunocompromised individuals.

Finally, it is worth considering whether individuals with prolonged SARS-CoV-2 shedding provide a crystal ball into future virus evolution. The E484A variant was identified in multiple immunosuppressed individuals before it appeared as a signature mutation in the Omicron Variant of Concern. Surveillance programs that concentrate on identifying prolonged infections and characterizing viral evolution in such cases may therefore provide important insights into the potential future directions of SARS-CoV-2 evolution.

## Materials and methods

### Ethics statement

Collection and testing of biological specimens and protected health information were performed in concordance with the University of Wisconsin IRB # 2021-0076 following informed consent from the patient.

### SARS-CoV-2 quantification

Diagnostic RT-qPCR was performed on nasopharyngeal swab specimens at Exact Sciences (Madison, WI), University of Wisconsin Hospitals and Clinics, or the Mayo Clinic for all timepoints except the one from ∼10 months post-diagnosis. On that timepoint, the patient was tested with a qualitative Panther Fusion assay from Hologic, Inc. (Marlborough, MA) for clinical care purposes (Supplemental Table 1).

To obtain a semi-quantitative measurement of the ∼10-month virus, we isolated viral RNA from an additional nasopharyngeal swab sample using the Total Nucleic Acid kit for the Maxwell RSC instrument (Promega, Madison, WI). We used the N1 qRT-PCR assay developed by the CDC and commercially available from IDT (Coralville, IA). The assay was run on a LightCycler 96 instrument (Roche, Indianapolis, IN) with the Taqman Fast Virus 1-step Master Mix enzyme (Thermo Fisher, Waltham, MA).

### SARS-CoV-2 culture

Vero E6/TMPRSS2 cells obtained from the Japanese Collection of Research Bioresources (JCRB) Cell Bank (1819) were grown in Dulbecco’s Modified Eagle Medium supplemented with 5% fetal calf serum, HEPES, amphotericin B, and gentamicin sulfate. The immunocompromised individual’s virus was isolated from the ∼10-months post-diagnosis nasopharyngeal swab sample cultured on Vero E6/TMPRSS2 cells. A prototypical 2020 SARS-CoV-2 control virus whose Spike protein sequence matched the ancestral Wuhan-1 reference except for D614G (SARS-CoV-2/UT-HP095-1N/ Human/2020/Tokyo) was described previously ^23,24^.

### Monoclonal antibody susceptibility assay

Monoclonal antibodies derived from the sequences of commercial therapeutic antibodies (Regeneron and Eli Lilly) were purchased from InvivoGen (Casirivimab, Imdevimab, Bamlanivimab, and Etesevimab). We diluted antibodies in cell culture media with 2-fold serial dilutions; final concentration of 10 µg/ml to 0.02 µg/ml in triplicate. Viruses were diluted in cell culture media and added to the diluted antibody series in wells of 96-well U-bottom plates to an adjusted titer of 100 plaque-forming units (PFU). Plates were incubated at 37ºC for 30 minutes. Culture supernatant was aspirated from Vero E6/TMPRSS2 cells plated in tissue culture 96-plates and replaced with the mixtures of serially diluted antibody and virus mixture (100µl/well) followed by incubation at 37°C with 5% CO2. The inhibitor concentration that prevented 99% cell death (IC99 value) at the given concentration (as determined by cytopathic effects visualized by a brightfield microscope) defines antibody potency.

### SARS-CoV-2 sequencing

We performed sequencing on a nasopharyngeal or nasal swab sample that was leftover after twelve of the immunocompromised individual’s PCR tests. All viral RNA extractions over the course of this study took place on the Maxwell RSC Viral Total Nucleic Acid Purification Kit (Promega, USA), used according to the manufacturer’s instructions.

To generate sequence data, we first sequenced viral cDNA libraries on an Oxford Nanopore (ONT) minION with the ARTIC v3 amplicon approach (https://www.protocols.io/view/covid-19-artic-v3-illumina-library-construction-an-bibtkann), but the previously documented amplicon dropout near the beginning of the Spike gene occurred in all of our samples. To improve breadth of coverage across the whole of the Spike gene, we re-sequenced ten of the samples on an ONT MinION using the MIDNIGHT protocol (https://www.protocols.io/view/sars-cov2-genome-sequencing-protocol-1200bp-amplic-bwyppfvn)25. To examine minor variant frequencies with maximum breadth of coverage, we later used the ARTIC v4.1 protocol to sequence an overlapping ten samples on an Illumina MiSeq (https://www.protocols.io/view/sars-cov-2-sequencing-on-illumina-miseq-using-arti-bfefjjbn).

Preparation for ARTIC v4.1 sequencing began with reverse transcription, for which we combined 11 µl of leftover viral isolate from ten of the samples with 4µl of SSIV Vilo Master Mix (Invitrogen, USA) and 5µl of DEPC water. We then incubated these pools through 25°C for 10 minutes, 50°C for 10 minutes, then 85°C for 5 minutes, with a final indefinite hold of 4°C. Next, we amplified 2.5µl of cDNA in two independent multiplexed PCR reactions with 12.5µl of Phusion High Fidelity Master Mix (New England BioLabs), 6µl of DEPC water and 4µl of ARTIC primer pools. After an initial 30 second denaturation at 98°C, we amplified the cDNA replicates over 30 cycles of 98°C for 30 seconds, 98°C for 15 seconds, and 65°C for 5 min, with an indefinite hold at 4°C to finish. We then combined and purified the amplicon pools with a 1:1 concentration of AMPure XP beads (Beckman Coulter, USA), and then quantified them using the Qubit hsDNA kit (Invitrogen, USA). For those samples being sequenced on the Illumina MiSeq (see Supplemental Table 1), we diluted all samples to 150ng in 60µl for use as inputs for the TruSeq DNA Nano Kit (Illumina, USA). The final, purified amplicon pools were then sequenced with the MiSeq reagent v3-600 Kit at 10pM with 1% PhiX. For those samples being sequenced with ARTIC on an ONT minION (see Supplemental Table 1), we mixed 12 µl of the purified DNA library, 37.5 µl of the ONT Sequencing Buffer II, and 25.5 µl of ONT Loading Solution, and loaded the mixture onto the MinION for sequencing. Additionally, our only sequence data from our timepoint at approximately 11 months post-diagnosis was sequenced on an Illumina MiSeq using an ARTIC v3 amplicon approach that was very similar to that described above, except that it uses tagmentation to fragment and barcode viral vDNA prior to sequencing (https://www.protocols.io/view/sars-cov-2-sequencing-on-illumina-miseq-using-arti-n92ld9w1xg5b/v1) ^26^.

To reverse transcribe the viral isolate in preparation for sequencing with the MIDNIGHT protocol, we combined 2 µl of LunaScript RT SuperMix (New England BioLabs, USA) with 8 µl of sample, and then incubated the pools at 25°C for 2 minutes, 55°C for 10 minutes, 95°C for 1 minute, and a final hold at 4°C. Next, we combined 2.5 μl of each incubated isolate together into one of two pools (pool A and pool B), both with 3.7 µl of nuclease-free water, 0.05 µl of MIDNIGHT primer pool, and 6.25 µl of Q5 HS Master Mix (New England BioLabs). We then amplified this mix with an initial 30 second denaturation at 98°C followed by 35 cycles of 98°C for 15 seconds, 61°C for 2 mins, and 65°C for 3 mins, with an indefinite 4°C hold to finish. 5 µl from each of these PCR product pools were then mixed with 2.5 μl of sample-specific barcode and 2.5ul of nuclease-free water, multiplexed, purified with 1X SPRI beads (Beckman Coulter, USA), and quantified with the Qubit dsDNA HS Assay Kit. Finally, a mixture of 12 µl of the purified DNA library, containing approximate 800 ng of DNA, along with 37.5 µl of the ONT Sequencing Buffer II, and 25.5 µl of ONT Loading Solution were loaded onto the ONT MinION Instrument for sequencing. The full MIDNIGHT protocol is available at https://www.protocols.io/view/sars-cov2-genome-sequencing-protocol-1200bp-amplic-bwyppfvn^25^.

### SARS-CoV-2 sequence analysis

#### Processing raw sequence data for consensus sequence analyses

The majority of analyses described here use consensus sequences, which we assembled for each sample using a workflow customized to run remotely with compute resources from the UW-Madison Center For High Throughput Computing (https://chtc.cs.wisc.edu/), and using the workload manager HTC Condor. Briefly, our SARS-CoV-2 sequence analysis pipeline base-calls reads with the Guppy basecaller. It then trims adapters and primers, maps to the SARS-CoV-2 Wuhan-1 sequence (GenBank: MN908947.3), and removes reads that map to more than one location. From there, consensus sequences are assembled with the requirement that each base is present in at least 50% of the downsampled reads. Any bases with a depth of less than 20 and/or an allele fraction under 50% reads are assigned an N. The pipeline then formats consensus sequences and metadata for upload to public sequence data repositories. In the separate workflow produced specifically for this study, we also call variants from MN908947.3 in these consensus sequences using iVar, which produces simpler outputs than the standard VCF, making them more straightforward to plot in R ^27^.

To visualize amplicon dropouts in these consensus sequences, we returned to the ONT MIDNIGHT reads in FASTQ format, which were originally used to generate consensus sequences. First, we mapped the reads to MN908947.3 using minimap2’s ‘map-ont’ preset, clipped reads down to MIDNIGHT amplicon pileups using SAMtools, and then used covtobed to produce BED-format tables of regions where read depth-of-coverage was below 20 ^28–30^. Finally, we used an R script to plot mutations in the consensus sequences through time, along with dropouts.

#### Processing Illumina reads for variant frequency analyses

While our sequence analysis pipeline is most refined for MIDNIGHT ONT data, it comes with the necessary caveat that ONT sequencing instruments have a relatively high error rate. This error rate is unproblematic when generating consensus sequences, but can also obscure low-frequency variants when examining intrahost variant frequencies. To overcome this obstacle, we sequenced leftover samples from ten timepoints on an Illumina MiSeq as described above. We then mapped these reads to MN908947.3 using minimap2’s ‘sr’ preset ^29^, clipped reads down to ARTIC amplicon pileups using SAMtools ^28^, and called variants in standard VCF format using the BBTools ‘callvariants.sh’ script ^31^. We then annotated protein effects onto these VCFs using snpEff ^32^. From there, variants that rose into consensus frequency after Bamlanivimab treatment were plotted using an R script.

#### Root-to-tip analysis

To examine the rate at which the patient’s virus accumulated mutations, and compare that rate with SARS-CoV-2 globally, we randomly subsampled approximately 5,000 SARS-CoV-2 sequences from NCBI GenBank. We then mapped the GenBank consensus sequences to MN908947.3 using minimap2, called variants from all sequence alignments using the BBTools ‘callvariants.sh’ script, and then used R to compute genetic distance from MN908947.3 and plot.

## Supporting information

Titles and Captions for Supplemental Figures and Tables

## Data Availability

All scripts, consensus sequence data, and sequencing read data used in the writing of this manuscript can be found at https://go.wisc.edu/89wtu8. Additional data from intermediate stages of our analysis are available upon reasonable request to the authors.

https://github.com/dholab/E484T-visualizations

https://go.wisc.edu/89wtu8

## Data Availability

All scripts, consensus sequence data, and sequencing read data used in the writing of this manuscript can be found at https://github.com/dholab/E484T-visualizations. Additional data from intermediate stages of our analysis are available upon reasonable request to the authors.

## Acknowledgements

We thank the teams at the Wisconsin State Laboratory of Hygiene and Wisconsin Division of Public Health for their work on the case and in the community writ large. We also thank Max Bobholz, William Vuyk, Corina Valencia, Wanting Wei, Siyang Peng and the whole sequencing team at the AIDS Vaccine Research Laboratory. This work was supported in part by the National Institutes of Allergy and Infectious Diseases Center for Research on Influenza Pathogenesis (HHSN272201400008C), the Center for Research on Influenza Pathogenesis and Transmission (CRIPT) (75N93021C00014), and by CDC Contract #75D30121C11060. Consensus sequence data was processed using the compute resources and assistance of the UW-Madison Center For High Throughput Computing (CHTC), which is supported by UW-Madison, the Advanced Computing Initiative, the Wisconsin Alumni Research Foundation, the Wisconsin Institutes for Discovery, and the National Science Foundation.

## Author contributions

G.K.M., K.M.B., K.K.R., T.C.F., and D.H.O. initiated the project and prepared the early stages of the manuscript. T.C.F., and D.H.O. further managed the project for its duration. N.R.M. designed analyses, produced figures and tables, and prepared the manuscript in its later stages. P.J.H. cultured the individual’s virus performed neutralization studies. L.A.H. and R.M. contributed sequence data and performed all associated laboratory work. A.P., K.J.A., M.C.L., and J.D.Y. managed treatment of the individual, provided case notes, and offered valuable feedback throughout the study. N.S., D.R.A., Y.Y., and M.F. provided critical review and insight at various stages in the project.

## Competing interests

We declare no conflicts of interest.

## Notes

### Competing Interest Statement

The authors have declared no competing interest.

### Summary of Updates

In this version, we add a visual comparison of the immunocompromised individual's persistently infecting virus with 5,000 SARS-CoV-2 sequences from GenBank.

