## Supplementary figures and images for "Evolution of a globally unique SARS-CoV-2 Spike E484T monoclonal antibody escape mutation in a persistently infected, immunocompromised individual"

### Supplemental Figure 1

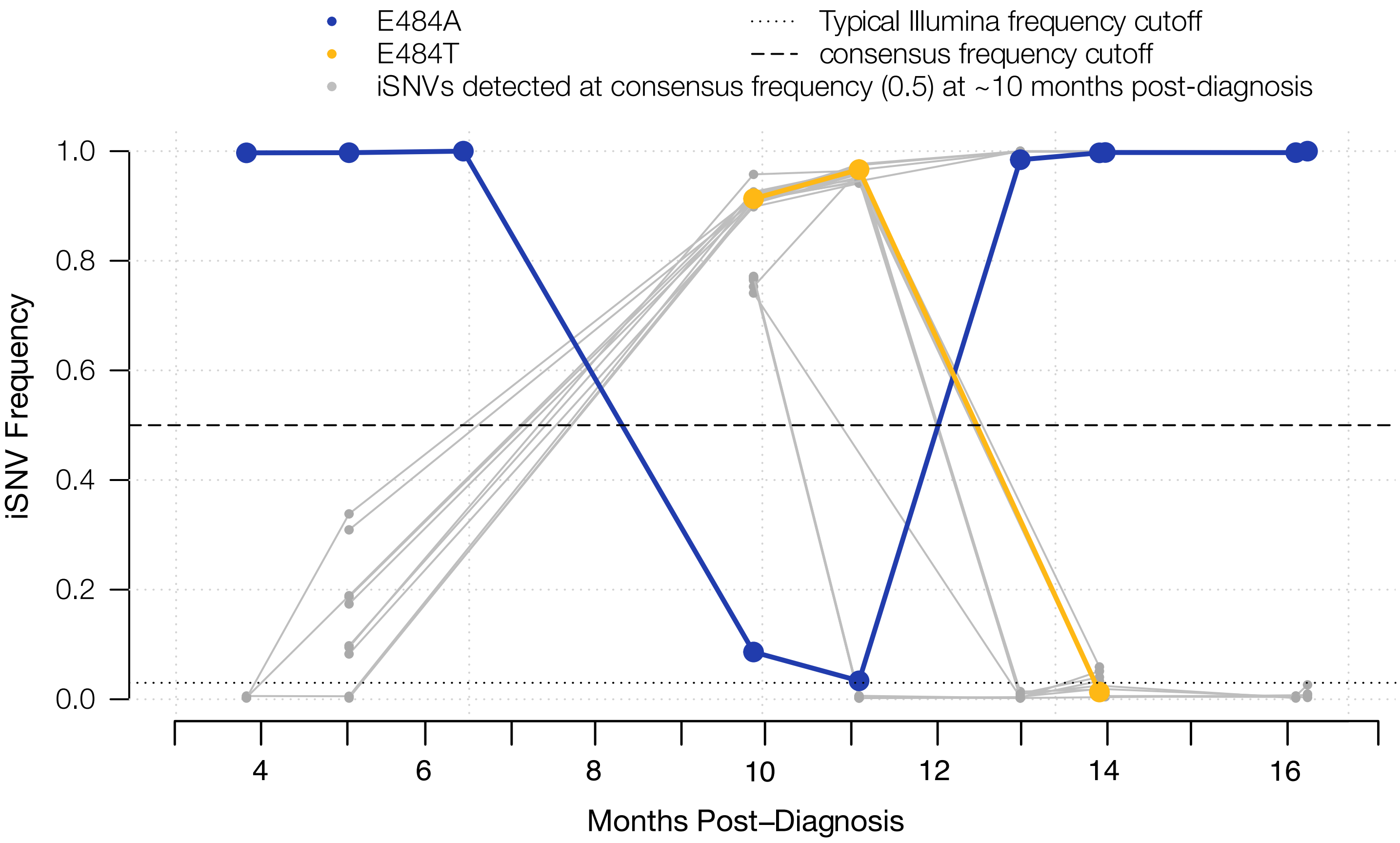

### Supplemental Figure 2

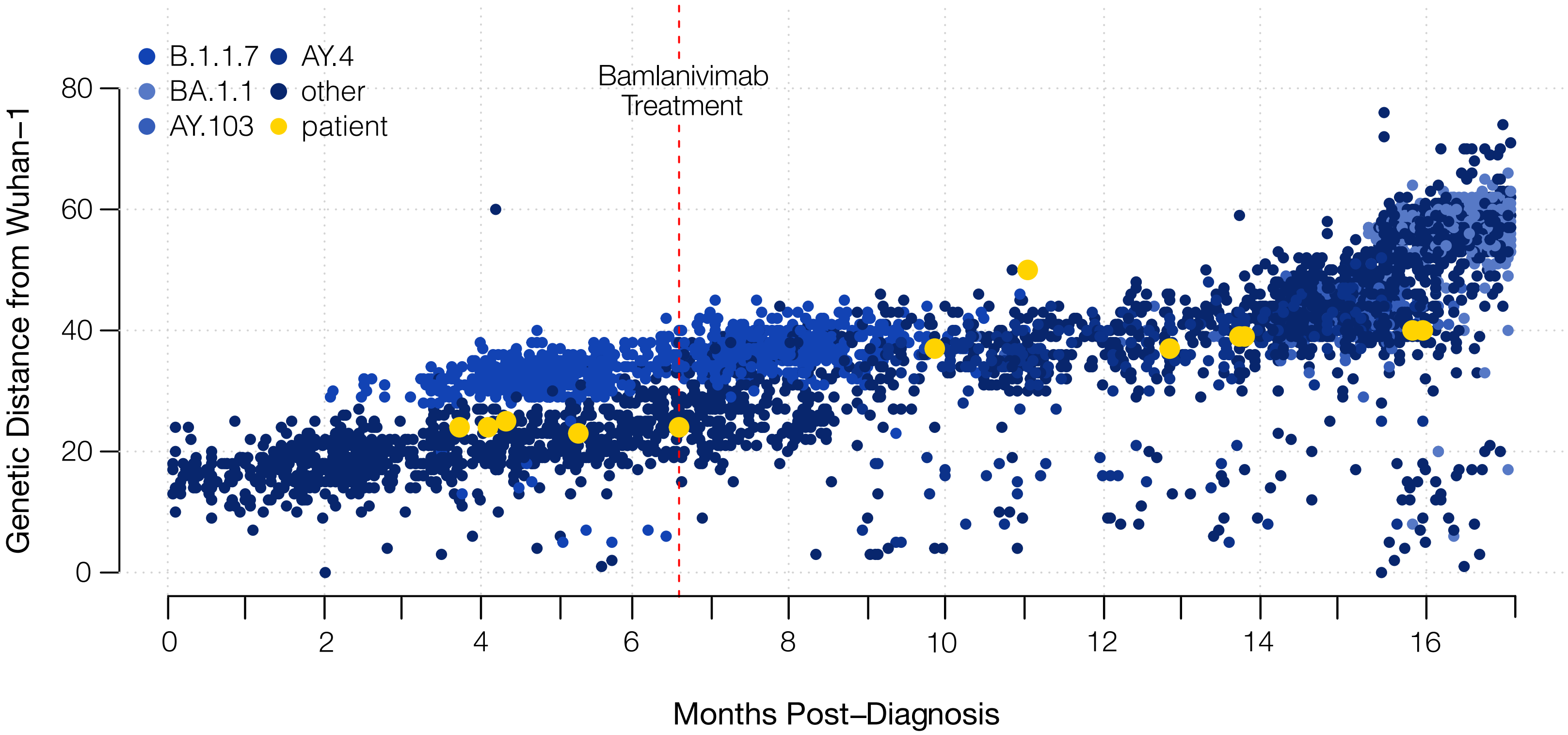
