## Supplementary material for "Evolution of a globally unique SARS-CoV-2 Spike E484T monoclonal antibody escape mutation in a persistently infected, immunocompromised individual": Titles and Captions for Supplemental Figures and Tables

#### Supplemental Figure 1.

Intrahost single-nucleotide variants (iSNVs) that reach ≥50% (consensus) frequency after Bamlanivumab monoclonal antibody treatment. Eleven of these iSNVs that reached consensus frequency after approximately 10 months of infection were present earlier in the infection, albeit at lower frequencies among sequencing reads (see Supplemental Table 2).

#### Supplemental Figure 2.

Root-to-tip analysis of chronic infection virus compared to a global subsample of 5,000 GenBank SARS-CoV-2 uploads. All points represent a sum of genetic differences between the sample consensus sequence and Wuhan-1 (GenBank: MN908947.3). Global SARS-CoV-2 samples accrue mutations according to a consistent clock rate, which is recapitulated by the chronic infection virus. A pronounced increase in mutations occurs after the Bamlanivumab monoclonal antibody treatment at approximately 6 months of infection.

### Supplemental Tables

#### Supplemental Table 1

Due to the collaborative nature of this project, virus in the immunocompromised individual's nasopharyngeal (NP) swab samples was quantified and sequenced using different platforms throughout the infection. All samples were sequenced with either the MIDNIGHT protocol on an Oxford Nanopore MinION, the ARTIC protocol on an Illumina MiSeq, or both.

#### Supplemental Table 2

25 Intrahost Single-Nucleotide Variants (iSNVs) that had risen above 0.5 frequency by 10 months post-diagnosis, with iSNV frequency among sequencing reads and depth-of-coverage compared between two library preparation methods. We detected eleven of these iSNVs at lower than consensus frequency in previous sequencing timepoints, whereas fourteen were first detected at approximately 10 months post-diagnosis. The nucleotide substitution that causes E484A, A-23013-C, is also included. Note that while G-23012-A causes Spike E484K in isolation, as it is listed in this table, the substitution caused Spike E484T in the patient's virus as a second-step from Spike E484A.
